# Age of menopause and dementia risk in 10,832 women from the Swedish Twin Registry

**DOI:** 10.1101/2024.11.13.24317223

**Authors:** Ursula G. Saelzler, Erin E. Sundermann, Janelle T. Foret, Margaret Gatz, Ida K. Karlsson, Matthew S. Panizzon

## Abstract

**INTRODUCTION:** An earlier age of menopause (AOM) is hypothesized to increase vulnerability to the neuropathological processes of dementia which begin in midlife.

**METHODS:** We tested this hypothesis in a sample of 10,832 women from the Swedish Twin Registry, stratified by menopause etiology. Survival models showed that a U-shaped association was present for women whose menopause occurred spontaneously. Sensitivity analyses conducted in hormone naïve, *APOE* ε4+ and AOM restricted subsamples showed largely analogous patterns of results.

**DISCUSSION:** Supporting conclusions from basic research, our results suggest that estrogens (proxied here by AOM) interact with several biological pathways mediating dementia disease processes. In line with trends in hormone research across the past century, our findings challenge the oversimplified ’more-is-better’ perspective on hormone exposure. Specifically, the non-linear association we observed between AOM and dementia risk points to the involvement of distinct and interacting biological mechanisms beyond just estrogen levels.

## 1 Background

In contrast to the steady decline of testosterone levels observed in men across the adult lifespan^1^, women experience a sharp drop in the production of estrogens by the ovaries at the time of menopause^2,3^. Estrogens play a pivotal role in neuro- and cardio-protective pathways; therefore, an earlier age of menopause (AOM) and thus the prolonged experience of a relative poverty of endogenous estrogens is hypothesized to increase a woman’s vulnerability to dementia-related pathophysiological processes that often begin already in midlife^4–13^.

Despite plausible mechanisms emerging from preclinical research, direct tests of this estrogen-poverty hypothesis have produced mixed results and attempts to meaningfully synthesize this literature have been severely hindered by the methodological heterogeneity of the primary studies. For example, a 2016 meta-analysis of 13 studies including 19,449 women found no difference in dementia risk between the youngest and oldest AOM groups^14^. Conversely, two subsequent meta-analyses, including 14 and 11 primary studies respectively, reported a pattern of increased dementia risk associated with “early” menopause (AOM < 45) and primary ovarian insufficiency (AOM < 40) ^15,16^. However, sensitivity analyses revealed that the exclusion of an exceptionally large cohort contributing roughly 99% of the participant level data to both analyses^17^, nullified the effect.

A woman’s AOM is defined by the age of her final menstrual period, provided amenorrhea has occurred for 12 consecutive months. AOM is inherently a continuous variable without biologically meaningful categories. Therefore, in addition to reducing statistical power and increasing the probability of false positives, the use of arbitrary categories, as was done in many of the previous investigations, limits statistical power and hinders between-study comparisons^18,19^.

Further, permanent amenorrhea can result from the spontaneous depletion of ovarian follicles (i.e., spontaneous or natural menopause), the removal of the ovaries via bilateral oophorectomy (i.e., induced or surgical menopause), or the removal of the uterus via hysterectomy. Despite producing analogous phenotypes, each of the three etiologies is associated with a distinct hormone profile^20,21^. Given that the change in estrogen levels is hypothesized to mediate the association between AOM and dementia risk, it is crucial to consider menopause etiology so that the results can be meaningfully interpreted in the context of plausible mechanisms. However, few studies collect sufficient gynecological surgery histories to characterize women’s menopause etiology. Similarly, the use of exogenous hormones, either through oral contraceptives (OC) or menopausal hormone therapy (MHT), has widespread impacts on the endocrine system, but lifetime history of hormone use is rarely available.

Given these noted limitations, we aim to contribute to the literature by examining associations between continuous AOM and dementia risk within a large sample of Swedish women with detailed reproductive health histories including previous gynecological surgeries and lifetime history of exogenous hormone use. This comprehensive data set enabled continuous analyses stratified by menopause etiology and sensitivity analyses conducted in a sample of exogenous hormone naïve women in order to understand the influence of menopause etiology and hormone use on the AOM and dementia connection.

## 2 Methods

Data were obtained from female individuals from the Swedish Twin Registry (STR) who participated in the Screening Across the Lifespan Twin (SALT) study between 1998 and 2002 (see Figure 1). Detailed information regarding the STR research infrastructure and SALT protocols can be found elsewhere^22,23^. Briefly, SALT aimed to screen for the most common complex diseases among twins enrolled in the STR born prior to 1959. Through a computer-assisted structured telephone interview, researchers screened participants for cognitive impairment and collected detailed self-reported health data including female-specific health factors such as AOM, births, history of gynecological surgeries, OC and MHT use.

**Figure 1.**
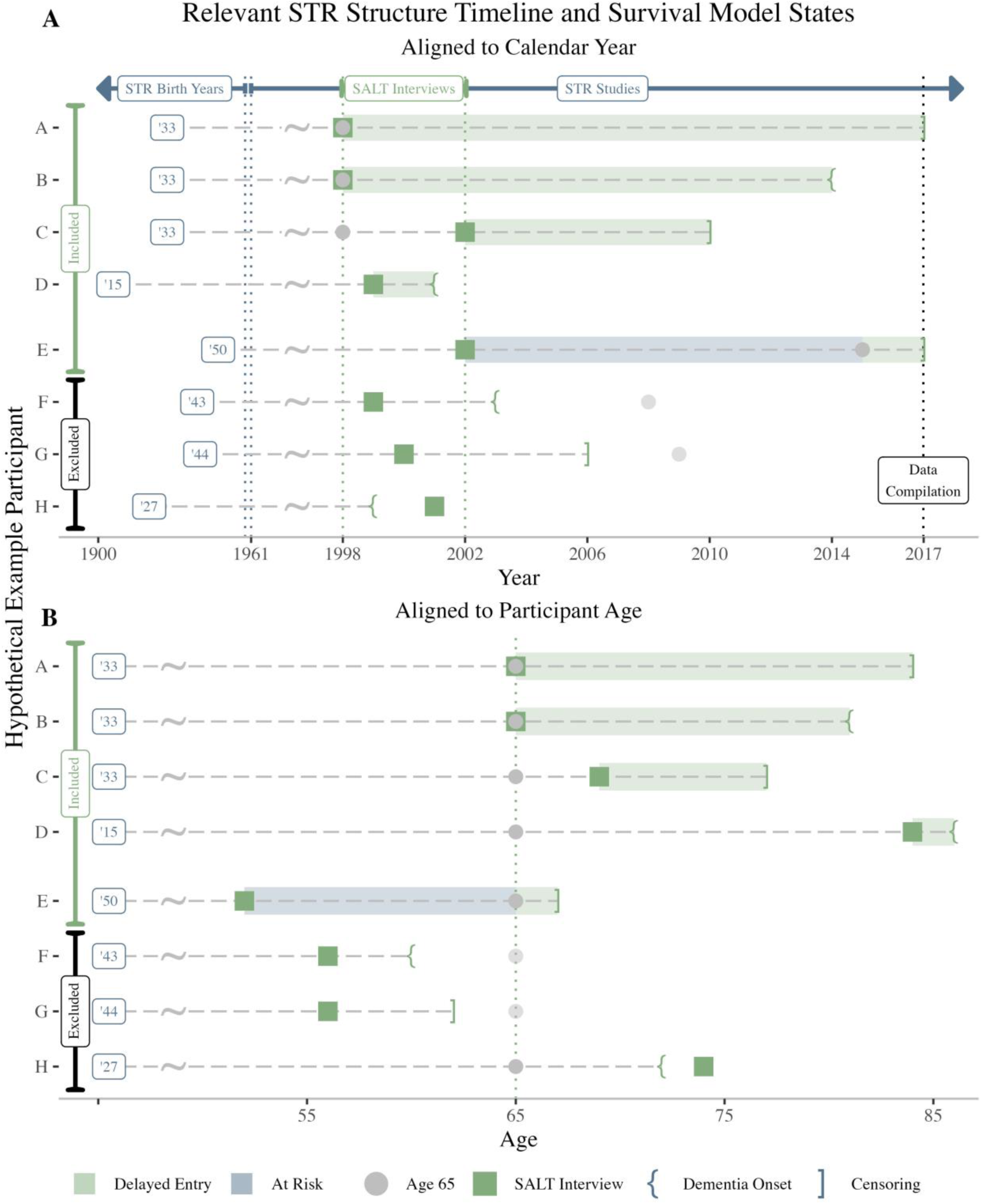
Graphical depiction of the Swedish Twin Registry (STR) research infrastructure, survival model states and hypothetical example individuals (HEIs) considered for inclusion current analyses. Panel A shows the HEIs aligned with the calendar year and Panel B reflects the HEIs aligned to age. HEIs A-D reflect typical entrants. That is, individuals who were 65 or older at the time of her Screening Across the Lifespan Twin (SALT) study interview and therefore enter the ’at risk’ for dementia set at the time of interview. HEI E reflects a delayed entrant. That is, an individual who was younger than 65 at the time of her SALT interview and therefore was delayed from entering the ’at risk’ set until age 65. HEIs A and E were administratively censored at the time of data compilation, meaning that no evidence of dementia was observed directly through an STR study and no dementia indicators appeared in any of the National Patient Registers on or prior to December 31, 2016. HEIs B and D showed evidence of dementia during follow up, and were therefore removed from the ’at risk’ set at the time of estimated dementia onset. HEI C was censored prior to data compilation and was therefore removed from the ’at risk’ set at the time of death or emigration. HEIs F-H reflect STR participants that completed at least some portion of the SALT interview but were excluded from the survival analyses. HEI F and G were excluded because they experienced an event prior to age 65 (the start of time). HEI H was excluded due to dementia onset prior to the SALT interview.

### 2.1 Data Acquisition and Model Parameters

#### 2.1.1 Menopause

##### 2.1.1.1 Etiology

If women reported 12 consecutive months of amenorrhea that did not result from medical treatment or gynecological surgery, they were classified as having experienced spontaneous menopause (SM). If women reported a history of unilateral or bilateral oophorectomy (removal of one or both of the ovaries) or saplingo-oophorectomy (removal of the ovaries and fallopian tube(s)) concurrent with or prior to spontaneous amenorrhea, they were classified as having experienced induced menopause (IM), regardless of whether a hysterectomy (removal of the uterus) was performed concurrently. Women who reported a hysterectomy (without oophorectomy) prior to spontaneous or induced menopause were excluded from analyses to ensure that the AOM demarcates a hormonal, rather than phenotypic change (i.e., the absence of menses). Women who were still in the reproductive or menopause transition stages at the time of the SALT interview (i.e., those still experiencing regular periods and those experiencing irregular periods with less than 12 consecutive months of amenorrhea, respectively) were classified as premenopausal and were excluded from the analyses.

##### 2.1.1.2 Age

Consistent with the Stages of Reproductive Aging Workshop + 10 staging ^24^, the spontaneous AOM was defined as women’s self-reported age of final menstrual period, provided that at least 12 consecutive months of amenorrhea occurred between the final menstrual period and the time of report. The self-reported age of oophorectomy was used to define the AOM within the IM sample.

#### 2.1.2 Dementia

##### 2.1.2.1 Diagnosis

###### 2.1.2.1.1 STR Screening and MCCC Diagnosis

The protocols for diagnosing dementia in STR have been described in detail elsewhere^25,26^. Briefly, individuals 65 and older were given a cognitive screening instrument as part of the SALT protocol. Individuals showing evidence of impairment and their co-twin were referred to an in-person clinical visit with a physician and psychologist which resulted in a differential dementia diagnosis through a multidisciplinary clinical consensus conference (MCCC). Where possible, the etiology of the dementia was ascertained and those presenting with dementia resulting from causes independent of aging (e.g., brain trauma, Multiple Sclerosis, hydrocephalus, Parkinson’s disease, Korsakoff’s syndrome etc.) were excluded.

###### 2.1.2.1.2 National Patient Registers

In addition to the screening of those over age 65 at the time of the SALT interview, the cognitive status of all participants was monitored through linkages with national patient registers until December 31, 2016. Individuals were given a dementia diagnosis if a) diagnostic codes associated with dementia appeared in in- or out-patient health records, b) individuals were prescribed drugs used to treat dementia, or c) if dementia was listed as a contributing factor to death^26^.

##### 2.1.2.2 Age of onset

For participants diagnosed with dementia via STR MCCC, an age of dementia onset was estimated through a combination of patient and informant interviews and medical record review^27^. For those participants diagnosed with dementia only through the National Patient Register, the age of onset was estimated using a linear model that leveraged the observed discrepancies between STR MCCC estimated onset and the age of register appearance among the 419 women with both sources of data and a MCCC estimated age of onset later than 65.

Congruent with a previous investigation^28^, the difference between the MCCC estimated age of dementia onset and the appearance of a dementia-associated diagnostic code or prescription record in a patient register averaged 4-5 years, and an average of 9 years elapsed between the MCCC age of onset and death.

If this model predicted an age of onset occurring after 65 but prior to the SALT interview, the age of onset was modified to be the midpoint between the SALT interview and the age of register indicator appearance (see Figure 2). As noted in section 2.1.2.1.1, cognitive screening was performed for all participants aged 65 and older and those demonstrating impairments were referred for in-person follow up. Therefore, if the individual was showing signs of dementia at the time of interview, a MCCC diagnosis would be available, and no register-based estimate would have been calculated. Without additional information, the mid-point provides the best estimate for this group.

**Figure 2.**
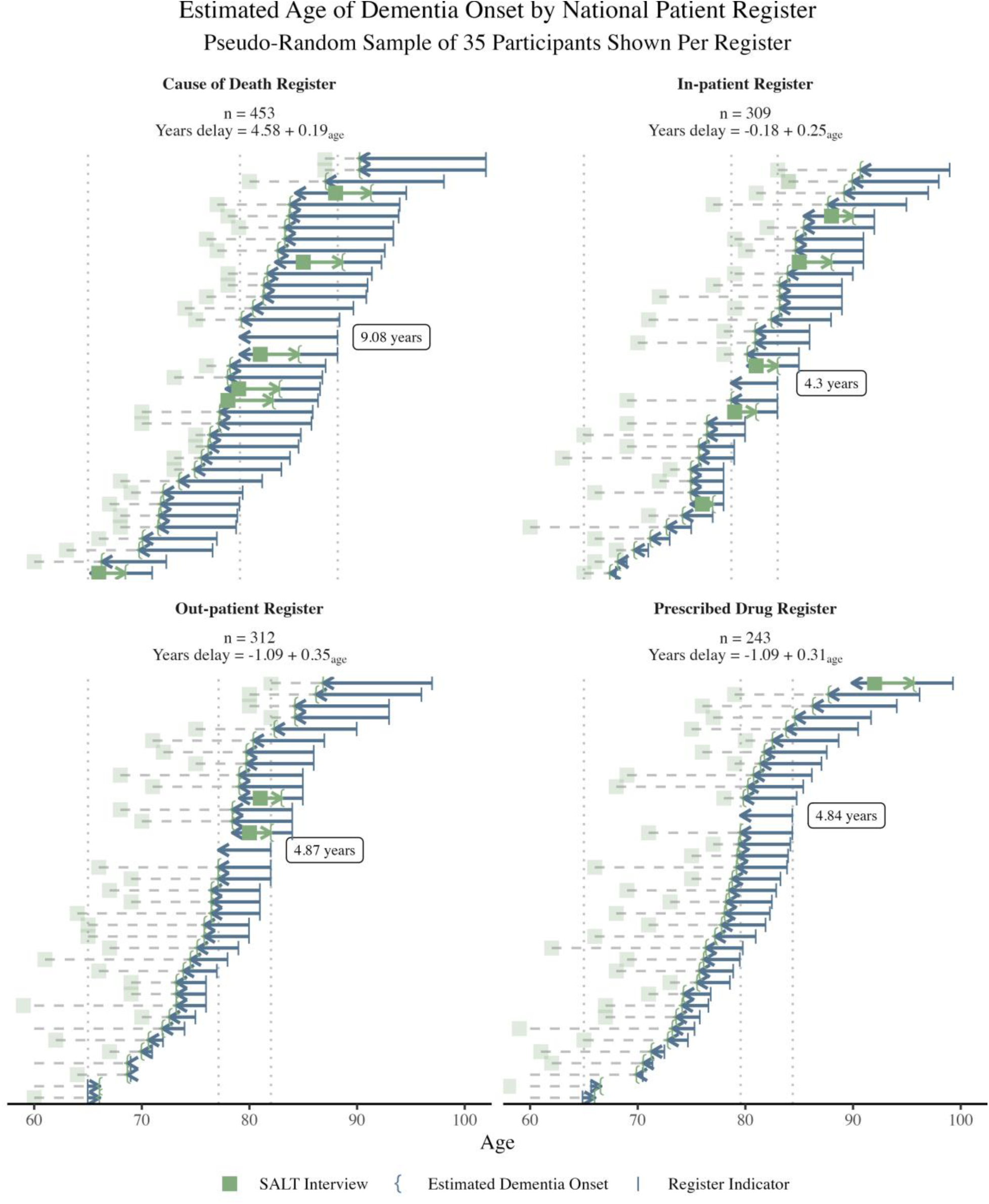
Estimation of age of dementia onset for female Screening Across the Lifespan Twin (SALT) participants with exclusively national patient register dementia indicators included in the primary analyses. Linear models were developed from a larger sample of all female STR participants with both a multidisciplinary clinical consensus conference (MCCC) dementia diagnosis and one or more register indicators of dementia in order to estimate the number of years that typically elapsed between a MCCC estimated time of dementia onset and the appearance of an indicator in a given national patient register. In all registers, age of register indicator appearance was positively associated with the estimated delay and was included as a covariate in all models, centered at age 65. The mathematical median delay per register is shown by the dotted vertical lines and indicated by the arrow to the left of the label. Cases where the linear model predicted an age of onset prior to the age of SALT interview, and therefore the mid-point estimation were used are indicated by the right facing arrows. Cases where the model-estimated age of dementia onset occurred after the SALT interview but prior to age 65 were excluded.

#### 2.1.3 Exclusionary Criteria

No missing data imputation was conducted. Therefore, only participants with complete data relating to the age and etiology of menopause, the diagnosis (or lack thereof) of dementia, an estimated age of onset when relevant, and all relevant covariates were included. All analyses were restricted to women with an AOM between 30 and 64 (inclusive). Women who reported a lifetime history of medical conditions known to impact menstrual cycles such as polycystic ovarian syndrome, or cancer were excluded. Finally, women for whom no follow-up data were available after age 65, who experienced onset of dementia prior to age 65 or prior to the SALT interview were also excluded.

### 2.2 Summary of Statistical Analyses

Data processing and analyses were conducted in R version 4.0.4 ^29^ utilizing the survival and survminer packages^30,31^. As visualized in Figure 1, age was used as the timescale for all CPHR models with age 65 demarcating the model start-of-time. Women whose SALT interview occurred after age 65 were treated as delayed entrants and were not included in the at-risk sample until her age of SALT interview. Dementia diagnosis was used as the model event and the estimated age of onset was used as the event time. Women that were not diagnosed with dementia during the observational period were censored (i.e., removed from the at-risk sample) at the age of death, emigration, or age on December 31, 2016. Observations were clustered by twin pair, ties were handled using Breslow’s exact method and the proportional hazards assumption was tested for all models using the method described by Grambsch and Therneau ^32^ via the *cox.zph* function.

For each question, an unadjusted and a covariate-adjusted model was applied to the relevant data. All covariate-adjusted models included three ordered factor covariates: the self-reported number of births (0, 1, 2, or 3+), lifetime educational attainment (primary or less, some or complete secondary, or post-secondary) and smoking history (ever regular smoker or never regular smoker).

#### 2.2.1 Primary Analyses

##### 2.2.1.1 Menopause etiology

To examine whether IM impacts dementia risk we applied model with menopause etiology as a binary variable (IM versus SM). AOM (centered at age 30) was included in the adjusted model.

##### 2.2.1.2 Linear and Quadratic AOM

To address our primary hypotheses regarding the association between continuous AOM and dementia, we fit two model sets each to the SM and IM samples. The first model imposed a linear association between AOM (centered at age 30) and dementia risk and the second imposed a quadratic association.

#### 2.2.2 Sensitivity Analyses

Planned sensitivity analyses applied the same series of models to three defined subsamples of women with SM: *APOE* ε4+, *APOE* ε4- and hormone naïve (i.e., never users of OC or HRT). Post-hoc sensitivity analyses were conducted in subsamples of the SM and IM groups restricted to those with an AOM < 61 to examine whether the pattern of results is influenced by the AOM range.

#### 2.2.3 Supplemental Analyses

##### 2.2.3.1 Primary Ovarian Insufficiency and Early Menopause

For comparison with existing literature that used a categorical definition of age at menopause, a second series of two model sets were fit to the SM sample. The first modeled primary ovarian insufficiency diagnosis (defined as AOM < 40) as a binary variable. The second modeled early menopause status (defined as AOM < 45) as a binary variable.

##### 2.2.3.2 Influence of Modeling Parameters

To provide real-world exploration of the impact of key model choices in the context of age-dependent disease outcomes, we modified the primary models (linear and quadratic associations between AOM and dementia) by 1) the use of time-on-study as timescale (rather than age, which we believe to be the appropriate timescale and therefore applied to all previous models), and/or 2) using “complete-cases” analyses (rather than delayed-entry) which necessitate the restriction of the sample to either: 1) those women who were under study prior to becoming “at risk” of the event, defined as age 65 here. That is, those women who were postmenopausal but not yet 65 at the time of her SALT interview. Or, 2) A sample restricted to those with equivalent opportunity to experience the factor of interest (post-menopausal status) prior to observation. That is, women who were 65 or older at the time of her SALT interview.

## 3 Results

A total of 10,832 women met the criteria for inclusion. As shown in Figure 3, the most common reasons for exclusion were being premenopausal at the time of the SALT interview or having an unknown AOM, accounting for 37% and 18% of excluded cases respectively.

**Figure 3.**
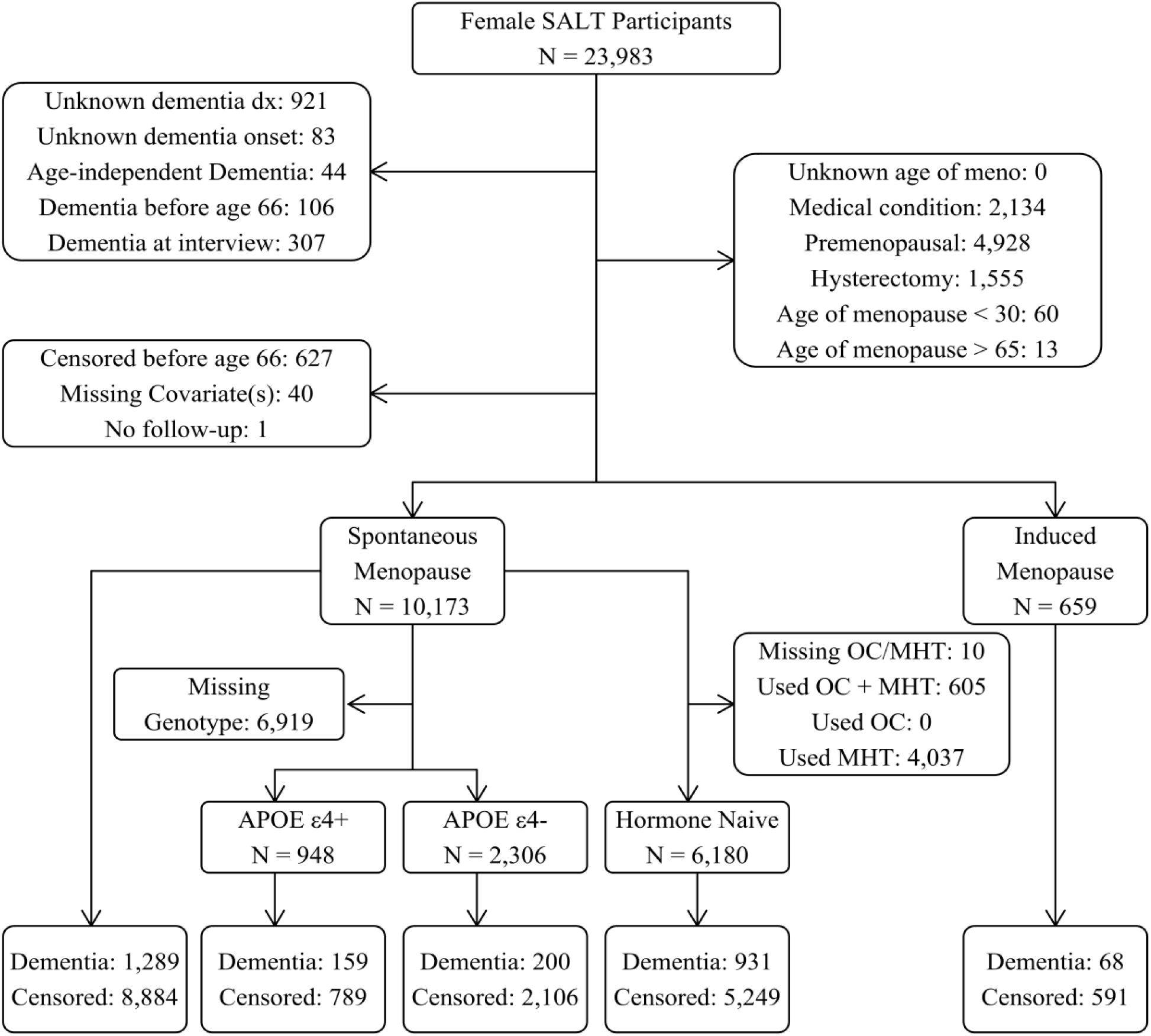
Participant inclusion flow chart for the primary and sensitivity analyses. OC: Oral contraceptive; MHT: Menopausal Hormone Therapy; SALT: Screening Across the Lifespan Twin Study; STR: Swedish Twin Registry.

Consistent with historical trends among Swedish women^33^ 94% of women experienced SM (see Table 1). Of the 10,173 women who experienced SM, 58% were considered exogenous hormone naïve (i.e., they reported never using exogenous hormones). Compared to hormone users, the hormone naïve participants were born earlier (average birth year 1933 versus 1939; see Supplemental Table 1), which is consistent with the introduction of OC in Sweden in the mid 1960s^34^ and increase in popularity of MHT during that same time^35^.

**Table 1.** Participant demographics for the spontaneous and induced menopause samples.

|  | Spontaneous Menopause |  |  |  | Induced Menopause |  |  |  |  |
| --- | --- | --- | --- | --- | --- | --- | --- | --- | --- |
|  | Total | Censored | Dementia | p<sup>a</sup> | Total | Censored | Dementia | p<sup>b</sup> | p<sup>c</sup> |
| n | 10,173 | 8,884 | 1,289 |  | 659 | 591 | 68 |  |  |
| Years Observed | 14.4 (4.7) | 15.5 (3.7) | 6.7 (3.8) | <0.001 | 14.5 (4.6) | 15.4 (3.8) | 6.8 (3.6) | <0.001 | 0.729 |
| Birth Year |  |  |  |  |  |  |  |  |  |
| Age | 1935.7 (9.7) | 1937.0 (9.4) | 1927.4 (7.5) | <0.001 | 1936.1 (9.5) | 1937.1 (9.3) | 1927.5 (5.9) | <0.001 | 0.401 |
| Menopause | 50.5 (4.0) | 50.5 (3.9) | 50.4 (4.5) | 0.155 | 47.2 (6.3) | 47.3 (6.3) | 46.1 (6.2) | 0.138 | <0.001 |
| SALT Interview | 63.9 (9.0) | 62.7 (8.7) | 71.6 (7.4) | <0.001 | 63.6 (8.9) | 62.6 (8.7) | 71.6 (5.8) | <0.001 | 0.410 |
| Event<sup>d</sup> | 78.3 (7.5) | 78.3 (7.8) | 78.3 (5.3) | 0.754 | 78.1 (7.5) | 78.0 (7.8) | 78.5 (4.7) | 0.636 | 0.439 |
| Births |  |  |  | <0.001 |  |  |  | 0.770 | 0.007 |
| 0 | 1,287 (12.7) | 1,099 (12.4) | 188 (14.6) |  | 103 (15.6) | 90 (15.2) | 13 (19.1) |  |  |
| 1 | 1,624 (16.0) | 1,383 (15.6) | 241 (18.7) |  | 124 (18.8) | 111 (18.8) | 13 (19.1) |  |  |
| 2 | 4,063 (39.9) | 3,645 (41.0) | 418 (32.4) |  | 256 (38.8) | 233 (39.4) | 23 (33.8) |  |  |
| 3+ | 3,199 (31.4) | 2,757 (31.0) | 442 (34.3) |  | 176 (26.7) | 157 (26.6) | 19 (27.9) |  |  |
| Education |  |  |  | <0.001 |  |  |  | 0.300 | 0.124 |
| Primary or less | 4,059 (39.9) | 3,323 (37.4) | 736 (57.1) |  | 281 (42.6) | 248 (42.0) | 33 (48.5) |  |  |
| Secondary<sup>e</sup> | 4,119 (40.5) | 3,683 (41.5) | 436 (33.8) |  | 269 (40.8) | 241 (40.8) | 28 (41.2) |  |  |
| Post-secondary | 1,995 (19.6) | 1,878 (21.1) | 117 (9.1) |  | 109 (16.5) | 102 (17.3) | 7 (10.3) |  |  |
| Ever Smoker<sup>f</sup> | 3,818 (37.5) | 3,460 (38.9) | 358 (27.8) | <0.001 | 278 (42.2) | 259 (43.8) | 19 (27.9) | 0.017 | 0.019 |
| *APOE ε4* | 948 (29.1) | 789 (27.3) | 159 (44.3) | <0.001 | 59 (26.0) | 50 (24.3) | 9 (42.9) | 0.112 | 0.350 |

|  | Spontaneous Menopause |  |  |  | Induced Menopause |  |  |  |  |
| --- | --- | --- | --- | --- | --- | --- | --- | --- | --- |
|  | Total | Censored | Dementia | p<sup>a</sup> | Total | Censored | Dementia | p<sup>b</sup> | p<sup>c</sup> |
| MHT Use | 4,268 (42.0) | 3,868 (43.6) | 400 (31.1) | <0.001 | 374 (56.8) | 350 (59.2) | 24 (35.3) | <0.001 | <0.001 |
| OC Use | 567 (6.2) | 545 (6.8) | 22 (1.8) | <0.001 | 38 (6.7) | 37 (7.4) | 1 (1.6) | 0.144 | 0.671 |

<sup>a</sup>Censored vs Dementia for the Spontaneous Menopause sample; <sup>b</sup>Censored vs Dementia for the Induced Menopause sample;<sup>c</sup>Spontaneous Menopause vs Induced Menopause samples; <sup>d</sup>Dementia or censoring; <sup>e</sup>Includes some secondary education; <sup>f</sup>Self-reported ever regularly smoking;
Note. OC: Oral contraceptive; MHT: Menopausal hormone therapy; SALT: Screening Across the Lifespan Study.

As anticipated given that evidence of dementia at the time of the SALT interview was an exclusion criterion, only 40 of the1,357 dementia cases had an onset date estimated by MCCC, the remaining estimated onset ages were based on patient register indicators. Of these estimates, 138 were adjusted to the midpoint as described in section 2.1.2.2.

### 3.1 Primary Analyses

There was some evidence for the violation of the proportional hazards assumption suggesting that the influence of ever regularly smoking on dementia risk is not consistent over time (see Supplemental Table 2). No other variables or global tests violated the proportional hazards assumption (ps > .05).

#### 3.1.1 Spontaneous Menopause

As shown in Table 3, Among women with SM, the linear model was a poor fit. Applying a quadratic model improved the fit, and this was unchanged in the adjusted models. Figure 4 shows the relative risk ratios derived from the quadratic model. The model indicates that a woman who experienced menopause at age 31 has more than 2.5 times the risk of developing dementia compared to a woman with an average age of menopause (around age 51). Similarly, a woman experiencing menopause at age 64 has almost 2 times greater risk of developing dementia compared to a woman with an average age of menopause across follow-up.

**Figure 4.**
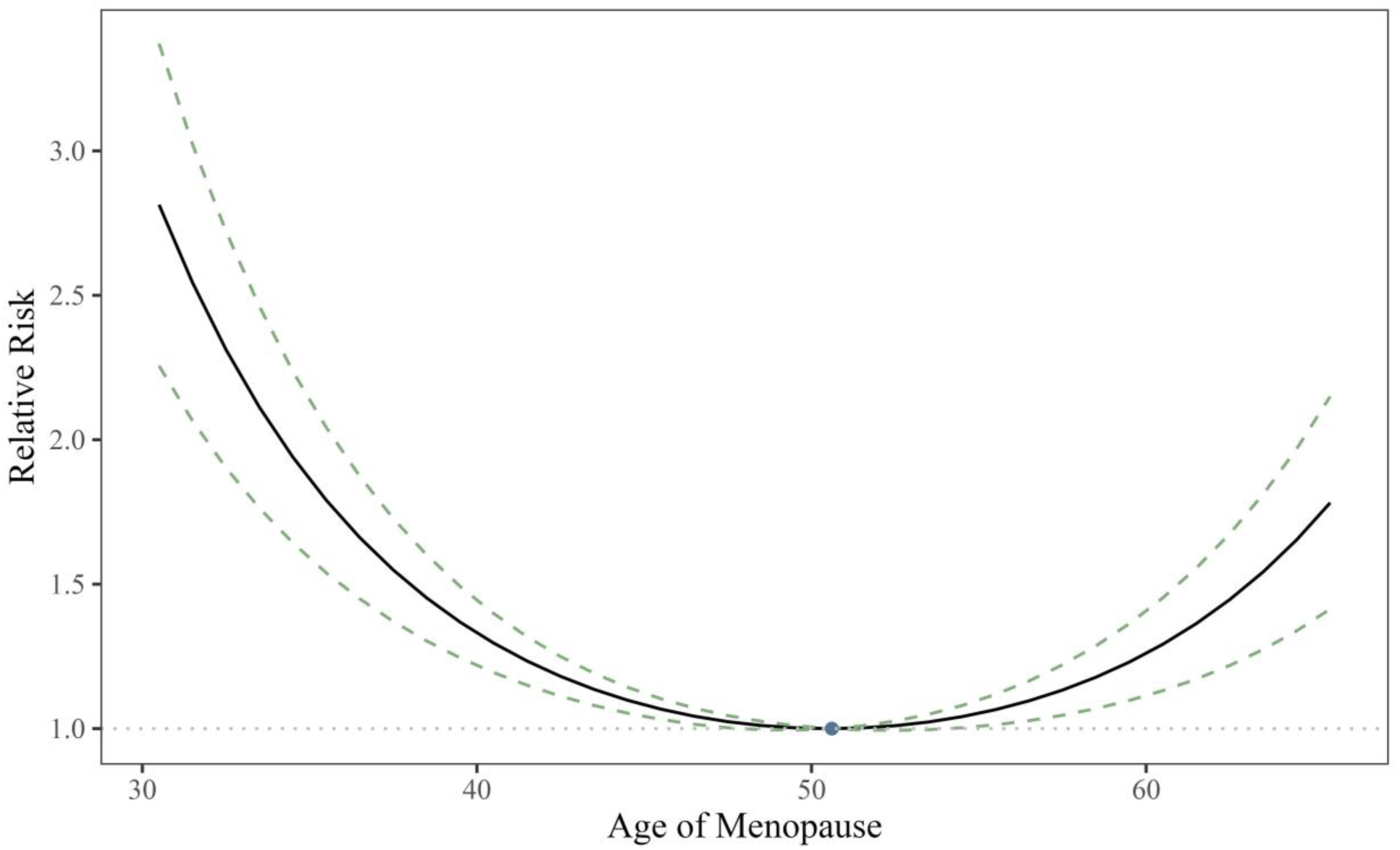
Relative risk of dementia by age of menopause (AOM) as predicted by the unadjusted quadratic model. Risk of dementia is shown relative to the predicted risk for a woman with the sample mean age of menopause (illustrated by the single point). The model indicates that a woman who experienced menopause at age 31 has more than 2.5 times the risk of developing dementia compared to a woman with an average age of menopause. Similarly, a woman experiencing menopause at age 64 has almost 2 times greater risk of developing dementia compared to a woman with an average age of menopause across follow-up.

#### 3.1.2 Induced Menopause

IM was not independently associated with dementia risk when covarying for AOM. Among women with IM, there was a negative linear association between the AOM and dementia risk. Specifically, each additional year of premenopausal status was associated with 5.8% lower dementia risk. There was no evidence of a quadratic association between AOM and dementia risk among women with IM (see Table 3).

### 3.2 Sensitivity Analyses

When stratified by *APOE* ε4 carrier status, the quadratic association between AOM and dementia risk only remained marginally significant for ε4 carriers (see Table 4). Restricting analyses to the hormone naïve subsample did not meaningfully change the pattern of results, nor did restricting analyses to the women who experienced SM prior to age 61 (see Supplemental Table 3).

### 3.3 Supplemental Analyses

#### 3.3.1 Primary Ovarian Insufficiency and Early Menopause

Early menopause (AOM < 45) was associated with an approximately 20% increase in dementia risk, while primary ovarian insufficiency (AOM < 40) was not significantly associated with dementia risk (see Table 2).

**Table 2.**
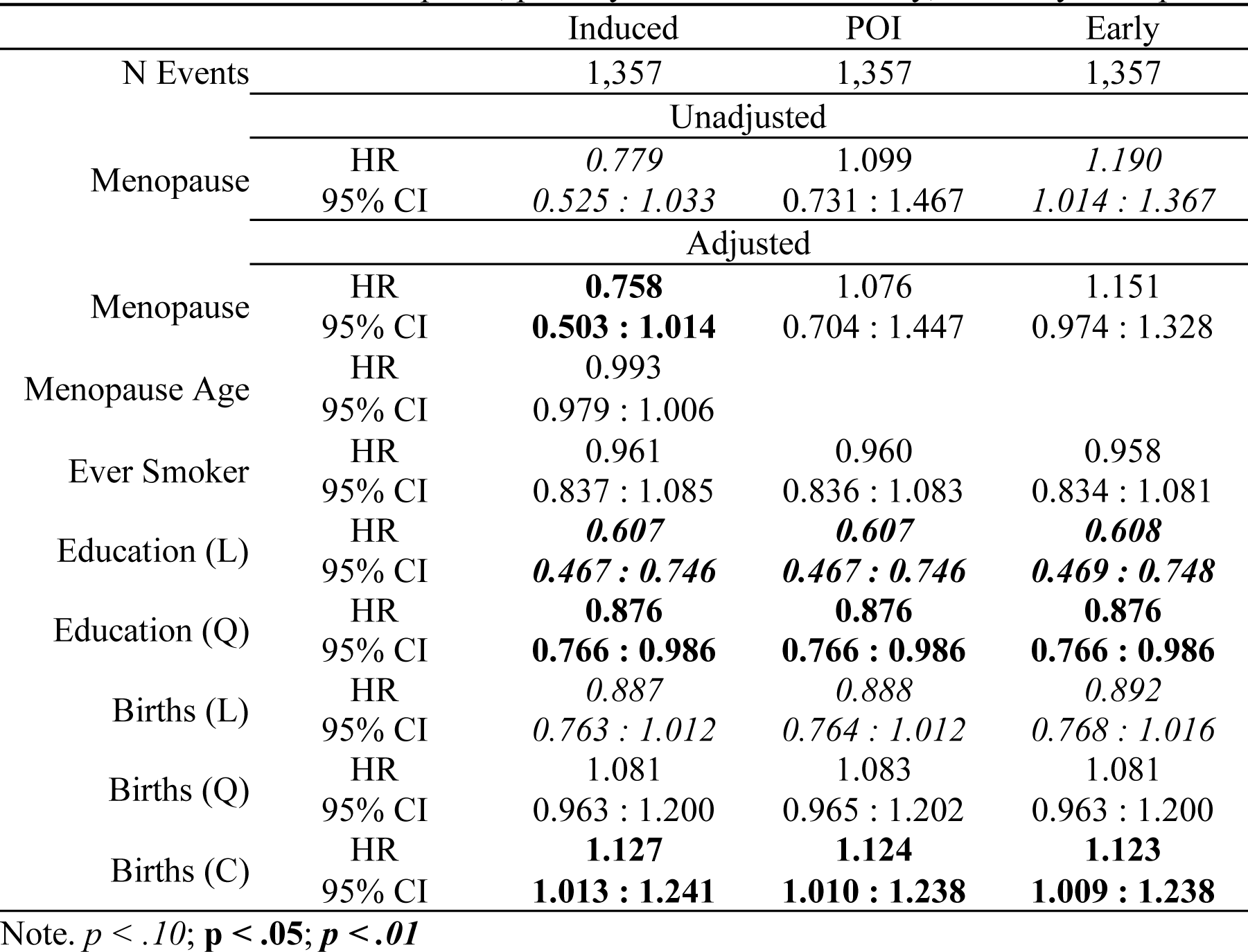
Results of the unadjusted and adjusted Cox proportional hazards model series examining the influence of induced menopause, primary ovarian insufficiency, and early menopause.

|  |  | Induced | POI | Early |
| --- | --- | --- | --- | --- |
| N Events |  | 1,357 | 1,357 | 1,357 |
|  |  | Unadjusted |  |  |
| Menopause | HR | 0.779 | 1.099 | 1.190 |
|  | 95% CI | 0.525 : 1.033 | 0.731 : 1.467 | 1.014 : 1.367 |
|  |  | Adjusted |  |  |
| Menopause | HR | **0.758** | 1.076 | 1.151 |
|  | 95% CI | **0.503 : 1.014** | 0.704 : 1.447 | 0.974 : 1.328 |
| Menopause Age | HR | 0.993 |  |  |
|  | 95% CI | 0.979 : 1.006 |  |  |
| Ever Smoker | HR | 0.961 | 0.960 | 0.958 |
|  | 95% CI | 0.837 : 1.085 | 0.836 : 1.083 | 0.834 : 1.081 |
| Education (L) | HR | **0.607** | **0.607** | **0.608** |
|  | 95% CI | **0.467 : 0.746** | **0.467 : 0.746** | **0.469 : 0.748** |
| Education (Q) | HR | **0.876** | **0.876** | **0.876** |
|  | 95% CI | **0.766 : 0.986** | **0.766 : 0.986** | **0.766 : 0.986** |
| Births (L) | HR | 0.887 | 0.888 | 0.892 |
|  | 95% CI | 0.763 : 1.012 | 0.764 : 1.012 | 0.768 : 1.016 |
| Births (Q) | HR | 1.081 | 1.083 | 1.081 |
|  | 95% CI | 0.963 : 1.200 | 0.965 : 1.202 | 0.963 : 1.200 |
| Births (C) | HR | **1.127** | **1.124** | **1.123** |
|  | 95% CI | **1.013 : 1.241** | **1.010 : 1.238** | **1.009 : 1.238** |

Note. $p < .10$ ; $p < .05$ ; $p < .01$

**Table 3.** Results of the unadjusted and adjusted Cox proportional hazards model series applied to the spontaneous and induced menopause samples.

|  |  | Spontaneous Menopause |  | Induced Menopause |  |
| --- | --- | --- | --- | --- | --- |
|  |  | Linear | Quadratic | Linear | Quadratic |
| N Events |  | 1,289 |  | 68 |  |
|  |  | Unadjusted |  |  |  |
| Menopause Age | HR | 0.992 | **0.900** | 0.965 | 1.002 |
|  | 95% CI | 0.978 : 1.007 | **0.830 : 0.969** | 0.928 : 1.003 | 0.845 : 1.158 |
| Menopause Age<sup>2</sup> | HR |  | **1.003** |  | 0.999 |
|  | 95% CI |  | **1.001 : 1.004** |  | 0.993 : 1.004 |
|  |  | Adjusted |  |  |  |
| Menopause Age | HR | 0.996 | **0.895** | **0.960** | 0.998 |
|  | 95% CI | 0.981 : 1.011 | **0.826 : 0.965** | **0.918 : 1.001** | 0.827 : 1.168 |
| Menopause Age<sup>2</sup> | HR |  | **1.003** |  | 0.999 |
|  | 95% CI |  | **1.001 : 1.005** |  | 0.993 : 1.005 |
| Ever Smoker | HR | 0.976 | 0.978 | 0.705 | 0.704 |
|  | 95% CI | 0.848 : 1.103 | 0.851 : 1.106 | 0.160 : 1.251 | 0.159 : 1.248 |
| Education (L) | HR | **0.592** | **0.589** | 0.971 | 0.995 |
|  | 95% CI | **0.448 : 0.735** | **0.445 : 0.733** | 0.360 : 1.582 | 0.380 : 1.610 |
| Education (Q) | HR | **0.883** | **0.880** | 0.743 | 0.756 |
|  | 95% CI | **0.769 : 0.996** | **0.767 : 0.993** | 0.269 : 1.217 | 0.275 : 1.237 |
| Births (L) | HR | **0.880** | **0.880** | 1.088 | 1.073 |
|  | 95% CI | **0.753 : 1.007** | **0.752 : 1.007** | 0.525 : 1.651 | 0.515 : 1.631 |
| Births (Q) | HR | 1.079 | 1.079 | 1.058 | 1.084 |
|  | 95% CI | 0.957 : 1.201 | 0.957 : 1.201 | 0.554 : 1.561 | 0.556 : 1.612 |
| Births (C) | HR | **1.145** | **1.143** | 0.828 | 0.819 |
|  | 95% CI | **1.028 : 1.262** | **1.026 : 1.260** | 0.347 : 1.309 | 0.338 : 1.300 |

Note. $p < .10$ ; $p < .05$ ; $p < .01$
The linear (L), quadratic (Q) and cubic (C) terms reflect the pattern of the relative change in hazard across the ordered categories but are not interpreted as typical hazard ratios.

**Table 4.**
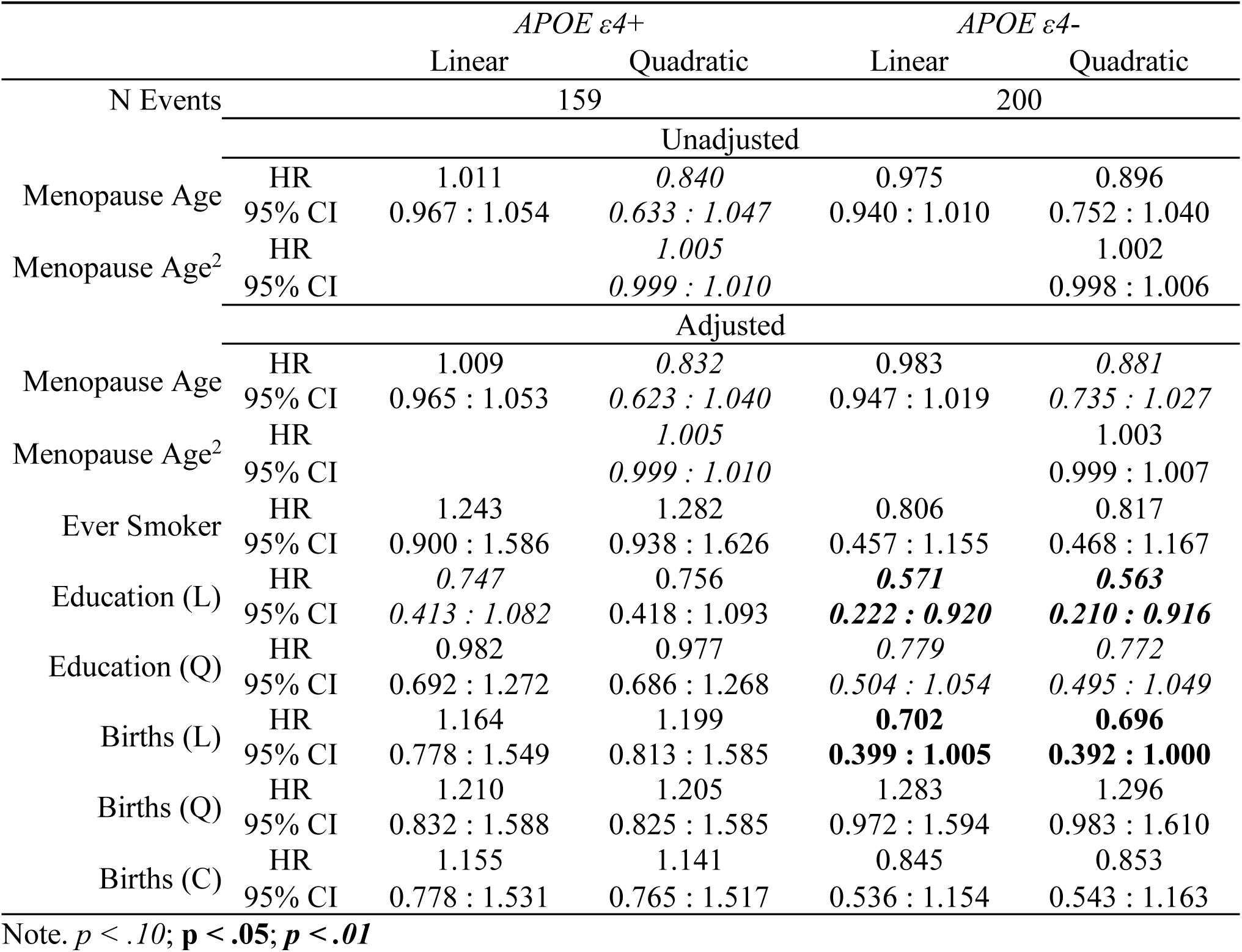
Results of the unadjusted and adjusted Cox proportional hazards model series applied to the APOE ε4+ and APOE ε4-samples using delayed entry with age as time scale.

|  |  | *APOE ε4+* |  | *APOE ε4-* |  |
| --- | --- | --- | --- | --- | --- |
|  |  | Linear | Quadratic | Linear | Quadratic |
| N Events |  | 159 |  | 200 |  |
|  |  | Unadjusted |  |  |  |
| Menopause Age | HR | 1.011 | *0.840* | 0.975 | 0.896 |
|  | 95% CI | 0.967 : 1.054 | *0.633 : 1.047* | 0.940 : 1.010 | 0.752 : 1.040 |
| Menopause Age<sup>2</sup> | HR |  | *1.005* |  | 1.002 |
|  | 95% CI |  | *0.999 : 1.010* |  | 0.998 : 1.006 |
|  |  | Adjusted |  |  |  |
| Menopause Age | HR | 1.009 | *0.832* | 0.983 | *0.881* |
|  | 95% CI | 0.965 : 1.053 | *0.623 : 1.040* | 0.947 : 1.019 | *0.735 : 1.027* |
| Menopause Age<sup>2</sup> | HR |  | *1.005* |  | 1.003 |
|  | 95% CI |  | *0.999 : 1.010* |  | 0.999 : 1.007 |
| Ever Smoker | HR | 1.243 | 1.282 | 0.806 | 0.817 |
|  | 95% CI | 0.900 : 1.586 | 0.938 : 1.626 | 0.457 : 1.155 | 0.468 : 1.167 |
| Education (L) | HR | *0.747* | 0.756 | **0.571** | **0.563** |
|  | 95% CI | *0.413 : 1.082* | 0.418 : 1.093 | **0.222 : 0.920** | **0.210 : 0.916** |
| Education (Q) | HR | 0.982 | 0.977 | *0.779* | *0.772* |
|  | 95% CI | 0.692 : 1.272 | 0.686 : 1.268 | *0.504 : 1.054* | *0.495 : 1.049* |
| Births (L) | HR | 1.164 | 1.199 | **0.702** | **0.696** |
|  | 95% CI | 0.778 : 1.549 | 0.813 : 1.585 | **0.399 : 1.005** | **0.392 : 1.000** |
| Births (Q) | HR | 1.210 | 1.205 | 1.283 | 1.296 |
|  | 95% CI | 0.832 : 1.588 | 0.825 : 1.585 | 0.972 : 1.594 | 0.983 : 1.610 |
| Births (C) | HR | 1.155 | 1.141 | 0.845 | 0.853 |
|  | 95% CI | 0.778 : 1.531 | 0.765 : 1.517 | 0.536 : 1.154 | 0.543 : 1.163 |

Note. $p < .10$ ; $p < .05$ ; $p < .01$

#### 3.3.2 Influence of Modeling Parameters

As shown in Supplemental Table 4 when time-on-study rather than age was used as the model time there was evidence that the proportional hazards assumption was violated, driven by interview age, for both the linear and quadratic models in both the SM and IM samples.

However, the CPHR model results were numerically similar to those observed in the primary analyses (see Supplemental Table 5).

Further, as anticipated, when analyses were conducted using complete cases rather than delayed entry, the sample size, and therefore the statistical power dropped (see Supplemental Table 6). Notably, just 4% of individuals who completed the SALT interview prior to age 66 received a dementia diagnosis during follow-up, compared to almost 25% of participants interviewed on or after age 65. This discrepancy is likely due to the increased risk of dementia with increasing age. Specifically, those with a younger interview age would also be followed at a younger age interval wherein they would be less at-risk for developing dementia than those interviewed at older ages. However, the quadratic association was only statistically significant in the sample interviewed prior to age 66 using time-on-study as model time (see Supplemental Table 7).

## 4 Discussion

Our analysis found that dementia risk increases with deviation from the typical age of menopause (AOM), regardless of direction among women with SM. Experiencing menopause 10 years earlier or later than the typical AOM was associated with a nearly 30% increase in dementia risk. While the estrogen hypothesis suggests a linear link between AOM and dementia risk, the quadratic association observed here suggests that estrogen interacts with multiple biological processes involved in the pathogenesis of dementia.

Consistent with prior findings, there was no difference in dementia risk between women with IM and those with SM^36^. Among women with IM, each additional reproductive year was linked to a 5% decrease in dementia risk.

However, we hesitate to interpret these findings in a biological context. Because the associated SALT question did not specify laterality, the IM classification was applied for either a bilateral or a unilateral oophorectomy despite the distinct hormone profiles associated with the surgeries^20,21^.

Interestingly, early menopause (AOM < 45) was linked to increased dementia risk, but primary ovarian insufficiency (AOM < 40) was not. However, in line with estimates from similar populations, just 2% of the sample experienced primary ovarian insufficiency, and of these women, just 31 also developed dementia during follow-up.

Notably, the quadratic association between AOM and dementia risk only remained marginally significant among *APOE* ε4 carriers. This is consistent with the evidence for interactive effects of estrogen with ε4 allele carriership ^37,38^, but should be interpreted with caution given the relatively small sample size.

### 4.1 Application of Survival Models to Cohort Studies

Cohort studies, like the STR, offer significant advantages for researching disease outcomes, including large sample sizes, comprehensive demographic and health data, and extended follow-up. However, cohort studies have several drawbacks and present a unique challenge for the application of CPHR models^39^.

#### 4.1.1 Reliance on patient registers

A limitation of the CPHR model is that participants cannot have experienced the event (dementia onset) before the model’s start (the later of age 65 or SALT interview). Only 3% of dementia onset events were defined by the MCCC, with the rest relying on patient register data. Register indicators, while highly specific, appeared for only 64.5% of MCCC-confirmed dementia cases.

The use of patient registers for dementia onset estimation presents a methodological challenge for survival models due to the delay between the onset of dementia symptoms and the point at which medical care (or death) results in a register-recorded indicator. This delay complicates survival modeling, which depends on meaningful and theoretically consistent event times. To address this, we utilized data from MCCC defined dementia cases within the STR to model the delay and estimate the true age of dementia onset based on the specific patient register and participant age.

#### 4.1.2 Reliance on self-report

AOM was self-reported. Previous investigations demonstrate that when recalling AOM, nearly 70% of women are accurate within 1-year, even after almost a decade ^40,41^. Further, even after almost two decades, 55% of women confirmed a previous report within 1-year^42^. However, both validity and reliability are reduced with increasing time since menopause^41,42^ while regression to the mean and the tendency to report an AOM of 50 increase^42^. Given that there was a median of 11.5 years between the self-reported AOM and the SALT interview, some inaccuracy in AOM exists.

Gynecological surgery history was also self-reported. A recent study found that while 80% of women reporting a hysterectomy had a corresponding medical record, one in three women reporting bilateral oophorectomy did not have a corresponding medical record^43^. Thus, the IM sample likely includes women with SM, potentially affecting power, though inclusion of SM women in the IM sample is unlikely.

#### 4.1.3 Relevant Sample Size

Like other regression models, the power of the CPHR models stems from the sample size. However, model power is based on the number of individuals experiencing the event, rather than total sample size. Although overly simplistic, it is generally accepted that a minimum of 10 events are needed per covariate for adequate power^44^. As shown in Table 1, just 68 women experienced both IM and dementia during follow-up, suggesting that the adjusted model is slightly underpowered for the ordered covariates despite a total sample size of over 1,000.

#### 4.1.4 Meaningful model start-of-time

Owing to their original application to intervention studies, study entry is often used as the model start-of-time. When applied to cohort studies however, this approach creates bias^45–48^ because only those surviving to the arbitrary time of study initiation (here, the SALT interviews conducted between 1998 and 2002) are included, while those who already experienced the event (dementia onset) are systematically excluded.

Age 65 was the optimal model start of time because it a) best accounts for the association between age and dementia risk, b) is the age that the cognitive screening was added to the SALT interview protocol, allowing for the confirmation of cognitive status at baseline for the majority of women, and c) is an age at which almost all women are post-menopausal meaning that all women experienced the “intervention” (menopause) prior to the model start-of-time

#### 4.1.5 Inability to capture time-varying covariates

Because the CPHR model requires that covariate values be known for individuals in the at-risk set at all model event times, which, in practice encompasses the entire timescale, factors of interest which are not stable across time cannot be meaningfully included in the models unless they are measured or can be reasonably estimated for all event times.

As an example, BMI was available at the time of SALT interview models. Following menopause, endogenous estrogen is primarily aromatized in adipose tissue ^49^, resulting in a positive association between post-menopausal adiposity and endogenous estrogen ^50,51^.

Therefore, within the estrogen hypothesis framework, higher adiposity should indirectly confer a neuroprotective effect, and there is some evidence of this effect ^52^. However, neither adiposity nor the association between BMI and fat mass are stable across time ^49^. Further, established associations between BMI and various health outcomes (including dementia risk) vary as a function of age^53^. Given that the age of SALT interview (and therefore the age of known BMI) ranged from 50 to 97, no cohesive interpretation of the construct reflected by the available measure of BMI could be determined.

### 4.2 Strengths and Limitations

Among the strengths of the current investigation is the STR’s development and early adoption of detailed demographic and women’s health history questionnaires. This investment enabled the statistical consideration of several key factors shown to impact both AOM and dementia risk^54,55^. Future studies utilizing combined interviews and medical record data are well poised to address current limitations^56,57^.

First, self-reported AOM is conceptualized here as a demarcation of a sharp and permanent decline in exogenous circulating estrogen levels. Although meaningfully true, the menopause transition occurs over a period of years and is characterized by large hormonal fluctuations followed by a consistent trend of decline until relative stability is reached and maintained through the post-menopausal period^24^. The nuance and individual differences in the transition could only be meaningfully captured by repeated hormone sampling beginning before the cessation of menses^58,59^. Such sampling would be logistically implausible and prohibitively expensive for most cohort studies.

Second, the influence of exogenous hormone use could not be fully examined. Although women provided specific OC and MHT formulations, even approaches which account for the relative differences in hormone dose (which decreased by an entire order of magnitude since their introduction compared to more modern formulations^60^), and the variance in the endocrine effects of the varying progestin (which can elicit opposing changes^60^) would not account for biologically meaningful factors such as duration and continuity of use or age of initiation.

In light of the critical window hypothesis which posits that MHT is beneficial if initiated shortly after menopause but detrimental if administered too long after the menopause transition^61,62^, as well as the close proximity of menopause to the potential initial accumulation of dementia pathology^9^, the delay between menopause is biologically interesting but could not be examined here. A further barrier to the meaningful consideration of associations between MHT and dementia risk in the context of the estrogen hypothesis is the primary use of MHT to alleviate menopausal symptoms, which are themselves frequently indicative of lower endogenous estrogen levels^63^.

Given the complexities of examining exogenous hormone use coupled with the relative infrequency of dementia, particularly amongst the women in the cohort born late enough to have exogenous hormones available, we elected to conduct sensitivity analyses in women that never used exogenous hormones, maximizing homogeneity.

### 4.3 Conclusion

Within a sample of over 10,000 postmenopausal women enrolled in the STR, we found that both early and late AOM conferred increased dementia risk in comparison to more typical AOM. These results are consistent with preclinical work showing interactions between estrogen and dementia processes. However, the mechanisms underlying the association between AOM and dementia risk have yet to be fully elucidated.

## Acknowledgements

We acknowledge The Swedish Twin Registry for access to data. The Swedish Twin Registry is managed by Karolinska Institutet and receives funding through the Swedish Research Council under the grant no 2017-00641.

## Author Approval

All authors have seen and approved the manuscript.

## Consent Statement

Consent was not necessary because this project only included secondary data analysis.

## Data Availability

Data can be requested from the Karolinska Institutet; https://ki.se/en/research/research-infrastructure-and-environments/core-facilities-for-research/the-swedish-twin-registry

## Abbreviations

AOM: Age of menopause
CPHR: Cox proportional hazards regression
IM: Induced menopause
OC: Oral contraceptives
MCCC: Multidisciplinary clinical consensus conference
MHT: Menopausal hormone therapy
SM: Spontaneous menopause
STR: Swedish Twin Registry

