## Supplement for "Age of menopause and dementia risk in 10,832 women from the Swedish Twin Registry"

Supplemental Table 1

Participant demographics for the hormone naive and exogenous hormone user samples

| | Hormone Naive | | | | Exogenous Hormone Users | | | | |
|---|---|---|---|---|---|---|---|---|---|
| | Total | Censored | Dementia | p[a] | Total | Censored | Dementia | p[b] | p[c] |
| n | 6,180 | 5,249 | 931 | | 4,642 | 4,218 | 424 | | |
| Years Observed | 13.7 (5.2) | 15.1 (4.2) | 6.1 (3.7) | <0.001 | 15.3 (3.7) | 16.1 (2.8) | 8.0 (3.6) | <0.001 | <0.001 |
| Birth Year | | | | | | | | | |
| Age | 1933.4 (10.0) | 1934.8 (9.8) | 1925.9 (7.3) | <0.001 | 1938.9 (8.2) | 1939.7 (7.9) | 1930.8 (6.7) | <0.001 | <0.001 |
| Menopause | 50.3 (4.1) | 50.3 (4.1) | 50.0 (4.6) | 0.060 | 50.3 (4.3) | 50.3 (4.2) | 50.4 (4.9) | 0.858 | 0.600 |
| SALT Interview | 66.0 (9.4) | 64.7 (9.2) | 73.1 (7.1) | <0.001 | 61.0 (7.5) | 60.2 (7.2) | 68.4 (6.5) | <0.001 | <0.001 |
| Event[d] | 79.7 (7.7) | 79.8 (8.0) | 79.2 (5.3) | 0.026 | 76.3 (6.8) | 76.3 (7.0) | 76.4 (4.8) | 0.713 | <0.001 |
| Births | | | | 0.002 | | | | <0.001 | <0.001 |
| 0 | 875 (14.2) | 733 (14.0) | 142 (15.3) | | 512 (11.0) | 454 (10.8) | 58 (13.7) | | |
| 1 | 1,052 (17.0) | 861 (16.4) | 191 (20.5) | | 694 (15.0) | 631 (15.0) | 63 (14.9) | | |
| 2 | 2,224 (36.0) | 1,932 (36.8) | 292 (31.4) | | 2,093 (45.1) | 1,945 (46.1) | 148 (34.9) | | |
| 3+ | 2,029 (32.8) | 1,723 (32.8) | 306 (32.9) | | 1,343 (28.9) | 1,188 (28.2) | 155 (36.6) | | |
| Education | | | | <0.001 | | | | <0.001 | <0.001 |
| Primary or less | 2,957 (47.8) | 2,386 (45.5) | 571 (61.3) | | 1,376 (29.6) | 1,180 (28.0) | 196 (46.2) | | |
| Secondary[e] | 2,361 (38.2) | 2,056 (39.2) | 305 (32.8) | | 2,026 (43.6) | 1,867 (44.3) | 159 (37.5) | | |
| Post-secondary | 862 (13.9) | 807 (15.4) | 55 (5.9) | | 1,240 (26.7) | 1,171 (27.8) | 69 (16.3) | | |
| Ever Smoker[f] | 2,147 (34.7) | 1,905 (36.3) | 242 (26.0) | <0.001 | 1,948 (42.0) | 1,813 (43.0) | 135 (31.8) | <0.001 | <0.001 |
| *APOE ε4* | 517 (28.2) | 406 (25.7) | 111 (44.0) | <0.001 | 490 (29.8) | 433 (28.5) | 57 (44.5) | <0.001 | 0.323 |
| PMHT Use | 0 (0.0) | 0 (0.0) | 0 (0.0) | NaN | 4,642 (100.0) | 4,218 (100.0) | 424 (100.0) | NaN | <0.001 |
| OC Use | 0 (0.0) | 0 (0.0) | 0 (0.0) | NaN | 605 (17.0) | 582 (18.1) | 23 (6.8) | <0.001 | <0.001 |

[a]Censored vs Dementia for the Hormone Naive sample; [b]Censored vs Dementia for the Exogenous Hormone Users sample; [c]Hormone Naive vs Exogenous Hormone Users samples; [d]Dementia or censoring; [e]Includes some secondary education; [f]Self-reported ever regularly smoking;

Note. OC: Oral contraceptive; PMHT: Post-menopausal hormone therapy; SALT: Screening Across the Lifespan Study.

Supplemental Table 2

Results of the cox.zph function for the primary analyses in the spontaneous and induced menopause samples using age as model time and delayed entry. The null hypothesis indicates no violation of the proportional hazards assumption.

| | Spontaneous Menopause | | | | | | Induced Menopause | | | | | |
|---|---|---|---|---|---|---|---|---|---|---|---|---|
| | Linear | | | Quadratic | | | Linear | | | Quadratic | | |
| | $\chi^2$ | df | *p* | $\chi^2$ | df | *p* | $\chi^2$ | df | *p* | $\chi^2$ | df | *p* |
| | | | | | | Unadjusted | | | | | | |
| Menopause Age | 0.568 | 1 | .451 | 0.767 | 1 | .381 | 1.599 | 1 | .206 | 1.763 | 1 | .184 |
| Menopause Age$^2$ | | | | 0.203 | 1 | .652 | | | | 1.773 | 1 | .183 |
| Global | 0.568 | 1 | .451 | 5.407 | 2 | .067 | 1.599 | 1 | .206 | 1.803 | 2 | .406 |
| | | | | | | Adjusted | | | | | | |
| Menopause Age | 0.286 | 1 | .593 | 0.406 | 1 | .524 | 1.711 | 1 | .191 | 1.860 | 1 | .173 |
| Menopause Age$^2$ | | | | 0.066 | 1 | .797 | | | | 1.887 | 1 | .170 |
| Ever Smoker | 8.517 | 1 | .004 | 8.518 | 1 | .004 | 0.097 | 1 | .756 | 0.106 | 1 | .745 |
| Education | 0.236 | 2 | .889 | 0.305 | 2 | .858 | 1.240 | 2 | .538 | 1.299 | 2 | .522 |
| Births | 0.855 | 3 | .836 | 0.791 | 3 | .852 | 1.717 | 3 | .633 | 1.761 | 3 | .624 |
| Global | 9.753 | 7 | .203 | 14.025 | 8 | .081 | 5.344 | 7 | .618 | 5.780 | 8 | .672 |

Note. *p < .10*; **p < .05**; ***p < .01***

Supplemental Table 3

Results of the unadjusted and adjusted Cox proportional hazards model series applied to hormone naive and AOM < 61 subsamples with age as model time

| | | Hormone Naive | | Spontaneous Menopause Age < 61 | | Induced Menopause Age < 61 | |
|---|---|---|---|---|---|---|---|
| | | Linear | Quadratic | Linear | Quadratic | Linear | Quadratic |
| N Events | | 931 | | 1,273 | | 67 | |
| | | Unadjusted | | | | | |
| Menopause Age | HR | 0.994 | **0.904** | *0.988* | **0.924** | **0.960** | 1.058 |
| | 95% CI | 0.977 : 1.011 | **0.824 : 0.983** | *0.973 : 1.002* | **0.849 : 0.999** | **0.924 : 0.996** | 0.903 : 1.213 |
| Menopause Age$^2$ | HR | | **1.003** | | *1.002* | | 0.997 |
| | 95% CI | | **1.000 : 1.005** | | *1.000 : 1.004* | | 0.991 : 1.002 |
| | | Adjusted | | | | | |
| Menopause Age | HR | 0.998 | ***0.900*** | 0.991 | **0.918** | **0.955** | 1.057 |
| | 95% CI | 0.981 : 1.015 | ***0.820 : 0.979*** | 0.977 : 1.006 | **0.842 : 0.995** | **0.915 : 0.995** | 0.891 : 1.224 |
| Menopause Age$^2$ | HR | | **1.003** | | **1.002** | | 0.997 |
| | 95% CI | | **1.001 : 1.005** | | **1.000 : 1.004** | | 0.991 : 1.002 |
| Ever Smoker | HR | 0.958 | 0.958 | 0.973 | 0.976 | 0.712 | 0.710 |
| | 95% CI | 0.804 : 1.111 | 0.805 : 1.112 | 0.845 : 1.101 | 0.847 : 1.104 | 0.164 : 1.261 | 0.163 : 1.258 |
| Education (L) | HR | ***0.571*** | ***0.567*** | ***0.586*** | ***0.584*** | 0.968 | 1.037 |
| | 95% CI | ***0.370 : 0.772*** | ***0.365 : 0.768*** | ***0.440 : 0.731*** | ***0.438 : 0.729*** | 0.355 : 1.582 | 0.423 : 1.651 |
| Education (Q) | HR | **0.823** | **0.820** | **0.884** | **0.882** | 0.758 | 0.793 |
| | 95% CI | **0.672 : 0.973** | **0.669 : 0.971** | **0.769 : 0.999** | **0.767 : 0.997** | 0.278 : 1.237 | 0.315 : 1.271 |
| Births (L) | HR | *0.862* | *0.862* | *0.885* | *0.884* | 1.108 | 1.068 |
| | 95% CI | *0.712 : 1.011* | *0.712 : 1.011* | *0.758 : 1.013* | *0.756 : 1.012* | 0.543 : 1.672 | 0.516 : 1.621 |
| Births (Q) | HR | 0.987 | 0.987 | 1.084 | 1.083 | 1.081 | 1.136 |
| | 95% CI | 0.846 : 1.129 | 0.845 : 1.128 | 0.962 : 1.207 | 0.961 : 1.206 | 0.571 : 1.591 | 0.618 : 1.654 |
| Births (C) | HR | *1.141* | *1.137* | **1.136** | **1.135** | 0.857 | 0.833 |
| | 95% CI | *1.005 : 1.277* | *1.001 : 1.273* | **1.019 : 1.254** | **1.017 : 1.253** | 0.373 : 1.340 | 0.347 : 1.319 |

Note. *p < .10*; **p < .05**; ***p < .01***

The linear (L), quadratic (Q) and cubic (C) terms reflect the pattern of the relative change in hazard across the ordered categories but are not interpreted as typical hazard ratios.

Supplemental Table 4

Results of the cox.zph function for the primary analyses in the spontaneous and induced menopause samples using time-on-study as model time and delayed entry. The null hypothesis indicates no violation of the proportional hazards assumption.

| | Spontaneous Menopause | | | | | | Induced Menopause | | | | | |
|---|---|---|---|---|---|---|---|---|---|---|---|---|
| | Linear | | | Quadratic | | | Linear | | | Quadratic | | |
| | $\chi^2$ | df | *p* | $\chi^2$ | df | *p* | $\chi^2$ | df | *p* | $\chi^2$ | df | *p* |
| | Unadjusted | | | | | | | | | | | |
| Menopause Age | 1.093 | 1 | .296 | 0.735 | 1 | .391 | 0.000 | 1 | .995 | 0.000 | 1 | .994 |
| Menopause Age$^2$ | | | | 0.610 | 1 | .435 | | | | 0.105 | 1 | .746 |
| Interview Age | 79.147 | 1 | .000 | 81.431 | 1 | .000 | 1.042 | 1 | .307 | 1.043 | 1 | .307 |
| Global | 79.230 | 2 | .000 | 81.562 | 3 | .000 | 1.042 | 2 | .594 | 2.715 | 3 | .438 |
| | Adjusted | | | | | | | | | | | |
| Menopause Age | 1.218 | 1 | .270 | 0.811 | 1 | .368 | 0.000 | 1 | 1.000 | 0.000 | 1 | 1.000 |
| Menopause Age$^2$ | | | | 0.678 | 1 | .410 | | | | 0.103 | 1 | .749 |
| Interview Age | 81.856 | 1 | .000 | 84.314 | 1 | .000 | 0.864 | 1 | .353 | 0.864 | 1 | .353 |
| Ever Smoker | 5.760 | 1 | .016 | 5.941 | 1 | .015 | 2.666 | 1 | .103 | 2.668 | 1 | .102 |
| Education | 0.682 | 2 | .711 | 0.754 | 2 | .686 | 4.584 | 2 | .101 | 4.580 | 2 | .101 |
| Births | 7.737 | 3 | .052 | 7.708 | 3 | .052 | 8.289 | 3 | .040 | 8.281 | 3 | .041 |
| Global | 85.097 | 8 | .000 | 87.376 | 9 | .000 | 17.663 | 8 | .024 | 18.146 | 9 | .034 |

Note. *p < .10*; **p < .05**; ***p < .01***

Supplemental Table 5
Results of the unadjusted and adjusted Cox proportional hazards model series applied to the spontaneous and induced menopause samples using time-on-study as model time

| | | Spontaneous Menopause | | Induced Menopause | |
|---|---|---|---|---|---|
| | | Linear | Quadratic | Linear | Quadratic |
| N Events | | 1,289 | | 68 | |
| | | Unadjusted | | | |
| Menopause Age | HR | 1.001 | ***0.898*** | *0.967* | 0.972 |
| | 95% CI | 0.986 : 1.016 | ***0.830 : 0.967*** | *0.927 : 1.007* | 0.808 : 1.135 |
| Interview Age | HR | ***1.084*** | ***1.085*** | ***1.093*** | ***1.093*** |
| | 95% CI | ***1.078 : 1.090*** | ***1.078 : 1.091*** | ***1.068 : 1.118*** | ***1.067 : 1.119*** |
| Menopause Age$^2$ | HR | | ***1.003*** | | 1.000 |
| | 95% CI | | ***1.001 : 1.005*** | | 0.994 : 1.006 |
| | | Adjusted | | | |
| Menopause Age | HR | 1.003 | ***0.893*** | *0.960* | 0.968 |
| | 95% CI | 0.988 : 1.017 | ***0.824 : 0.963*** | *0.918 : 1.003* | 0.783 : 1.152 |
| Interview Age | HR | ***1.081*** | ***1.082*** | ***1.097*** | ***1.097*** |
| | 95% CI | ***1.075 : 1.088*** | ***1.075 : 1.088*** | ***1.070 : 1.125*** | ***1.069 : 1.125*** |
| Menopause Age$^2$ | HR | | ***1.003*** | | 1.000 |
| | 95% CI | | ***1.001 : 1.005*** | | 0.993 : 1.006 |
| Ever Smoker | HR | 1.095 | 1.102 | 0.711 | 0.710 |
| | 95% CI | 0.968 : 1.223 | 0.974 : 1.229 | 0.162 : 1.260 | 0.161 : 1.260 |
| Education (L) | HR | ***0.682*** | ***0.678*** | 1.135 | 1.143 |
| | 95% CI | ***0.538 : 0.826*** | ***0.534 : 0.822*** | 0.508 : 1.763 | 0.499 : 1.786 |
| Education (Q) | HR | **0.891** | **0.890** | 0.721 | 0.724 |
| | 95% CI | **0.779 : 1.004** | **0.777 : 1.002** | 0.254 : 1.188 | 0.239 : 1.208 |
| Births (L) | HR | 0.987 | 0.987 | 1.092 | 1.090 |
| | 95% CI | 0.860 : 1.115 | 0.859 : 1.114 | 0.544 : 1.640 | 0.543 : 1.638 |
| Births (Q) | HR | 1.028 | 1.029 | 0.991 | 0.995 |
| | 95% CI | 0.906 : 1.150 | 0.907 : 1.151 | 0.494 : 1.489 | 0.473 : 1.517 |
| Births (C) | HR | 1.090 | 1.085 | 0.852 | 0.850 |
| | 95% CI | 0.972 : 1.207 | 0.967 : 1.203 | 0.377 : 1.326 | 0.375 : 1.325 |

Note. *p < .10*; **p < .05**; ***p < .01***
The linear (L), quadratic (Q) and cubic (C) terms reflect the pattern of the relative change in hazard across the ordered categories but are not interpreted as typical hazard ratios.

Supplemental Table 6

Participant demographics for the SALT Interview < 66 and SALT Interview > 65 complete cases samples

| | Interview Age < 65 | | | | Interview Age ≥ 65 | | | | |
|---|---|---|---|---|---|---|---|---|---|
| | Total | Censored | Dementia | p[a] | Total | Censored | Dementia | p[b] | p[c] |
| n | 5,833 | 5,619 | 214 | | 4,340 | 3,265 | 1,075 | | |
| Years Observed | 16.3 (2.2) | 16.5 (1.9) | 11.2 (2.8) | <0.001 | 11.9 (5.9) | 13.8 (5.1) | 5.8 (3.3) | <0.001 | <0.001 |
| Birth Year | | | | | | | | | |
| Age | 1942.8 (4.2) | 1942.9 (4.1) | 1938.9 (2.9) | <0.001 | 1926.3 (6.3) | 1926.7 (6.4) | 1925.2 (5.9) | <0.001 | <0.001 |
| Menopause | 50.7 (3.6) | 50.7 (3.6) | 51.1 (4.3) | 0.157 | 50.2 (4.3) | 50.2 (4.3) | 50.2 (4.5) | 0.752 | <0.001 |
| Interview | 57.2 (3.4) | 57.1 (3.4) | 60.5 (2.7) | <0.001 | 72.8 (6.2) | 72.4 (6.2) | 73.9 (5.8) | <0.001 | <0.001 |
| Event[d] | 73.6 (4.1) | 73.6 (4.1) | 71.7 (2.6) | <0.001 | 84.6 (6.3) | 86.2 (5.9) | 79.7 (4.7) | <0.001 | <0.001 |
| Births | | | | 0.076 | | | | 0.637 | <0.001 |
| 0 | 627 (10.7) | 603 (10.7) | 24 (11.2) | | 660 (15.2) | 496 (15.2) | 164 (15.3) | | |
| 1 | 844 (14.5) | 809 (14.4) | 35 (16.4) | | 780 (18.0) | 574 (17.6) | 206 (19.2) | | |
| 2 | 2,645 (45.3) | 2,566 (45.7) | 79 (36.9) | | 1,418 (32.7) | 1,079 (33.0) | 339 (31.5) | | |
| 3+ | 1,717 (29.4) | 1,641 (29.2) | 76 (35.5) | | 1,482 (34.1) | 1,116 (34.2) | 366 (34.0) | | |
| Education | | | | <0.001 | | | | <0.001 | <0.001 |
| Primary of less | 1,644 (28.2) | 1,552 (27.6) | 92 (43.0) | | 2,415 (55.6) | 1,771 (54.2) | 644 (59.9) | | |
| Secondary[e] | 2,704 (46.4) | 2,612 (46.5) | 92 (43.0) | | 1,415 (32.6) | 1,071 (32.8) | 344 (32.0) | | |
| Post-secondary | 1,485 (25.5) | 1,455 (25.9) | 30 (14.0) | | 510 (11.8) | 423 (13.0) | 87 (8.1) | | |
| Ever Smoker[f] | 2,694 (46.2) | 2,591 (46.1) | 103 (48.1) | 0.609 | 1,124 (25.9) | 869 (26.6) | 255 (23.7) | 0.066 | <0.001 |
| *APOE ε4* | 594 (30.2) | 559 (29.3) | 35 (58.3) | <0.001 | 354 (27.6) | 230 (23.4) | 124 (41.5) | <0.001 | 0.122 |
| HRT Use | 3,033 (52.0) | 2,926 (52.1) | 107 (50.0) | 0.595 | 1,235 (28.5) | 942 (28.9) | 293 (27.3) | 0.334 | <0.001 |
| OC Use | 508 (9.9) | 498 (10.1) | 10 (5.2) | 0.034 | 59 (1.5) | 47 (1.6) | 12 (1.2) | 0.483 | <0.001 |

[a]Censored vs Dementia for the Interview Age < 65 sample; [b]Censored vs Dementia for the Interview Age ≥ 65 sample; [c]Interview Age < 65 vs Interview Age ≥ 65 samples; [d]Dementia or censoring; [e]Includes some secondary education; [f]Self-reported ever regularly smoking;

Note. OC: Oral contraceptive; PMHT: Post-menopausal hormone therapy; SALT: Screening Across the Lifespan Study.

Supplemental Table 7

Results of the unadjusted and adjusted Cox proportional hazards model series applied to the the SALT Interview < 66 and SALT Interview > 65 complete cases samples using age as model time

| | | Interview Age < 66 | | Interview Age > 65 | |
|---|---|---|---|---|---|
| | | Linear | Quadratic | Linear | Quadratic |
| N Events | | 214 | | 1,075 | |
| | | Unadjusted | | | |
| Menopause Age | HR | 0.989 | ***0.799*** | 1.000 | *0.931* |
| | 95% CI | 0.948 : 1.030 | ***0.642 : 0.955*** | 0.985 : 1.016 | *0.856 : 1.007* |
| Interview Age | HR | ***1.278*** | ***1.274*** | | |
| | 95% CI | ***1.232 : 1.324*** | ***1.228 : 1.320*** | | |
| Menopause Age$^2$ | HR | | ***1.005*** | | *1.002* |
| | 95% CI | | ***1.001 : 1.009*** | | *1.000 : 1.004* |
| | | Adjusted | | | |
| Menopause Age | HR | 0.997 | ***0.808*** | 1.002 | **0.925** |
| | 95% CI | 0.956 : 1.039 | ***0.650 : 0.967*** | 0.987 : 1.018 | **0.848 : 1.002** |
| Interview Age | HR | ***1.279*** | ***1.275*** | | |
| | 95% CI | ***1.232 : 1.326*** | ***1.228 : 1.322*** | | |
| Menopause Age$^2$ | HR | | ***1.005*** | | **1.002** |
| | 95% CI | | ***1.001 : 1.009*** | | **1.000 : 1.004** |
| Ever Smoker | HR | **1.415** | **1.421** | 0.968 | 0.969 |
| | 95% CI | **1.143 : 1.687** | **1.149 : 1.693** | 0.819 : 1.116 | 0.820 : 1.117 |
| Education (L) | HR | ***0.645*** | ***0.647*** | ***0.646*** | ***0.642*** |
| | 95% CI | ***0.341 : 0.949*** | ***0.343 : 0.950*** | ***0.480 : 0.812*** | ***0.476 : 0.809*** |
| Education (Q) | HR | 0.831 | 0.830 | *0.892* | *0.889* |
| | 95% CI | 0.587 : 1.075 | 0.586 : 1.073 | *0.762 : 1.022* | *0.759 : 1.019* |
| Births (L) | HR | 0.934 | 0.936 | 0.897 | 0.897 |
| | 95% CI | 0.603 : 1.264 | 0.603 : 1.269 | 0.759 : 1.036 | 0.759 : 1.036 |
| Births (Q) | HR | 1.079 | 1.061 | 1.040 | 1.041 |
| | 95% CI | 0.770 : 1.388 | 0.750 : 1.373 | 0.906 : 1.173 | 0.908 : 1.175 |
| Births (C) | HR | *1.325* | *1.335* | 1.070 | 1.068 |
| | 95% CI | *1.026 : 1.624* | *1.037 : 1.634* | 0.941 : 1.198 | 0.939 : 1.196 |

Note. *p < .10*; **p < .05**; ***p < .01***

The linear (L), quadratic (Q) and cubic (C) terms reflect the pattern of the relative change in hazard across the ordered categories but are not interpreted as typical hazard ratios.
